# Impact of Lifting School Masking Requirements on Incidence of COVID-19 among Staff and Students in Greater-Boston Area School Districts: A Difference-in-Differences Analysis

**DOI:** 10.1101/2022.08.09.22278385

**Authors:** Tori L. Cowger, Jaylen Clarke, Eleanor J. Murray, Sarimer M. Sánchez, Mary T. Bassett, Bisola O. Ojikutu, Natalia Linos, Kathryn T. Hall

## Abstract

**Background:** In February 2022, following the rescinding of a Massachusetts statewide school masking mandate, only two cities (Boston and neighboring Chelsea) out of 79 school districts in the greater-Boston area, maintained masking requirements in K-12 schools. This provided an opportunity to examine the impact of removing masking on COVID-19 case rates among students and staff in the public-school setting.

**Methods:** We used difference-in-differences for staggered policy adoption to compare incidence of COVID-19 cases among students and staff in greater-Boston area school districts that lifted masking requirements to those that had not yet lifted masking requirements during the 2021-2022 school year.

**Results:** Before the statewide school masking policy was lifted, there was no statistically significant difference in case rate trajectories between school districts. However, weekly and cumulative case rates were significantly higher in students and staff in school districts that removed masking requirements, compared to districts that had not yet lifted requirements. We estimate that lifting of school masking requirements was associated with an additional 44.9 (95% CI: 32.6, 57.1) COVID-19 cases per 1,000 students and staff over the 15 weeks since the lifting of the statewide school masking requirement, representing nearly 30% of all cases observed in schools during that time. School districts that sustained masking requirements for longer periods tended to have older school buildings in poorer condition, more crowded classrooms, higher proportion of low income and English learning students and students with disabilities, and a higher proportion of Black and Latinx students and staff.

**Conclusions:** Masking is a relatively low-cost but effective intervention that can protect students and staff from substantial illness and loss of in-person days in school. Despite compelling evidence that masking significantly reduces the spread of SARS-CoV-2, political will and public adherence to masking has waned. Our study confirms that universal masking requirements can benefit all students and staff, and therefore represents an important strategy to mitigate the impacts of structural racism, ensure health equity, and to avoid potential deepening of educational inequities.

## INTRODUCTION

The direct and indirect impacts of the COVID-19 pandemic on children, their families, and surrounding communities have been substantial. Through February 2022, more children and adolescents in the U.S. have been infected with SARS-CoV-2 than any other age group.^1^ Mass infection during the Omicron wave was especially consequential for younger age groups. During December 2021 - February 2022, approximately one-third of all children and adolescents in the U.S. were newly infected with SARS-CoV-2, and peak hospital admissions for COVID-19 were at least 3 times as high as any other period during the pandemic.^1,2^ While the risk of severe COVID-19 is markedly lower compared with adults, children with COVID-19 are at risk for severe acute complications, including Multisystem Inflammatory Syndrome (MIS-C) as well as persistent long-term sequelae (i.e., long COVID/post-COVID conditions).^3–18^ Layered on these health impacts are substantial impacts to children’s social environments. More than 140,000 children and adolescents in the U.S. were estimated to have lost a parent or caregiver to COVID-19 through June 2021.^19^ In addition, COVID-19 has caused substantial interruptions in school settings, including staffing shortages, school closures, and missed school days, deepening existing educational inequities.^20,21^

Importantly, the impacts of COVID-19 have been disproportionately borne by groups made vulnerable by historic and contemporary systems of oppression, including structural racism and settler colonialism.^22–25^ Black, Indigenous, and Latinx children and adolescents were more likely to experience severe COVID-19 outcomes, death of parents/caregivers, worsening mental health, and educational disruptions, among other outcomes, compared to their white counterparts.^3,19,21,26–29^ These manifestations of structural racism underscore the urgent need to prioritize health equity in COVID-19 policies and programs that impact children and adolescents.

Schools represent an important venue for policies and interventions that minimize COVID-19’s impacts on students and staff. Even prior to the COVID-19 pandemic, schools were not uniformly health-promoting environments, shaped by environmental racism and chronic underinvestment. Historic and contemporary policies and practices including redlining, land theft, disinvestment and gentrification have eroded tax bases and shaped the quality of public school infrastructure and associated environmental hazards.^23,30–35^ These varying school conditions, including crowded classrooms, exposure to toxins and pests, and poor air quality due to outdated or absent HVAC/ventilation systems have left school districts differentially-equipped to respond to COVID-19, with harms concentrated among low income and Black, Latinx, and Indigenous communities.^30,36,37^

Alongside improved ventilation, vaccination, testing, and social supports to minimize the secondary impacts of COVID-19, masking represents an important piece of a layered mitigation strategy in school settings.^38–41^ A growing body of evidence supports the effectiveness of universal masking requirements in reducing SARS-CoV-2 transmission both in community and school settings.^42–48^ During the emergence of the Delta variant prior to the start of the 2021-2022 school year, CDC issued guidance recommending “… universal indoor masking by all students, staff, teachers, and visitors to K-12 schools, regardless of vaccination status”.^49^ However, on February 25, 2022, CDC released updated guidance that limited masking recommendations in public indoor settings (including K-12 schools) to counties with high COVID-19 Community Levels – a CDC-defined metric largely determined by COVID-19 hospitalizations.^50^ Following this updated guidance, many statewide policies requiring masks in both community settings and schools were lifted, largely shifting policy decisions to the local level.^51^

Massachusetts (MA) was one of only 18 states plus DC with statewide school masking requirements at some point during the 2021-2022 school year.^52^ The Massachusetts Department of Elementary and Secondary Education (DESE) lifted the statewide school masking requirement on February 28, 2022, joining a number of other states, from California to New Jersey, that similarly removed mask mandates in schools around the same time.^52^ With statewide school masking orders no longer in place and newly revised CDC guidance for public indoor settings, many MA school districts lifted masking requirements immediately, several sustained masking requirements for several weeks following the rescindment of the statewide order, and two districts – Boston and neighboring Chelsea Public Schools – maintained masking requirements through June 2022. The staggered lifting of masking requirements across Boston-area school districts presents a unique opportunity to evaluate the impact of school masking policies, during a period of highly transmissible SARS-CoV-2 variants and a rapidly changing COVID-19 policy environment.

The goal of this study was to examine the impact of staggered lifting of school masking requirements on the incidence of COVID-19 among staff and students in MA school districts, and to describe potential impacts of policy choices for health equity. Specifically, we aimed to (1) compare weekly incidence of COVID-19 in school districts that lifted masking requirements to districts where masking requirements had not yet been lifted; (2) estimate the difference in COVID-19 incidence among students and staff attributable to lifting mask protections (i.e., excess risk and population attributable fraction); and (3) compare school districts characteristics (e.g., staff/student sociodemographics, school building conditions, etc.) for districts that chose to sustain masking policies for longer periods of time to those that lifted masking requirements earlier.

## METHODS

### Data Sources & Definitions

We used publicly-available, school district-level data on COVID-19 cases, enrollment, and staffing for the 2021-2022 school year from DESE.^53,54^ Each Thursday, school districts are required to report COVID-19 cases among students and staff for the prior 7 days (Thursday-Wednesday). In addition to required case reporting throughout our study period, DESE offered state-sponsored testing programs options, including pooled and symptomatic testing and option introduced in mid-January for state-provided weekly take-home rapid antigen tests for schools that opt-in.^55,56^

For included school districts, we manually gathered data on dates of school and citywide masking policies from school district websites and/or local news sources as available. In addition, we obtained data on COVID-19 indicators in the surrounding communities (case rates, percent test positivity, testing rate) from the Massachusetts Department of Public Health (DPH) used for covariate adjustment in sensitivity analyses described below. Finally, for descriptive analyses, we extracted information on school district characteristics, including demographic information for students and staff, enrollment of DESE-defined selected populations (e.g., low-income students, English learners, students with disabilities, etc.) from DESE,^54^ and information on school building condition and learning environment from the 2016 School Survey (most recent available data) from the Massachusetts School Building Authority (MSBA).^57^

### Exposure and Outcomes

The primary exposure in this study was whether a school district maintained or lifted masking requirements in each reporting week. Under the statewide school masking requirement in place through February 28, 2022, all schools had masking requirements in place at the start of our study. A school district was considered to have lifted their masking requirement if their policy was rescinded prior to the first day of the reporting week (i.e., if the masking requirement was lifted part way through the reporting week, that week was classified as having masking requirements sustained).

Our primary outcome of interest was weekly reported rates of COVID-19 among staff and students. We reported rates as the number of COVID-19 cases per 1,000 staff and students (considered together), and separately among staff and students.

### Inclusion and Exclusion Criteria

For this analysis, we considered the n=79 school districts within the greater-Boston metro area contained within the U.S. Census Bureau-defined Boston-Cambridge-Newton New England City and Town Area (NECTA) division after excluding Charter and Vocational/Technical school districts (**Figure S1**). Of these, we excluded n=7 school districts with unreliable or missing testing data for more than 5 weeks of the study period (**Supplementary Appendix, Methods**). Our final sample included n=72 school districts representing n=294,084 students and n=46,530 staff over the 40 calendar weeks of the 2021-2022 school year through June 15, 2022 (**Table S1**).

### Statistical Analysis

We conducted a difference-in-differences analysis with staggered implementation to compare the weekly incidence of COVID-19 in school districts that lifted mask requirements compared to school districts where mask requirements had not yet been lifted.^58–61^ In this analysis, we estimated the weekly and cumulative impact over 15 weeks of removing mask requirements on reported COVID-19 cases in schools that removed masking requirements (i.e., average treatment effect among the treated).

### Sensitivity analyses

We conducted various sensitivity analyses to ensure our results were robust to changes in model specifications, data cleaning steps, and inclusion/exclusion criteria. Additional details can be found in the **Supplementary Appendix**. Briefly, we varied data cleaning procedures (raw data vs. corrections for 2-week reporting periods and corrections for non-reporting as zeroes), control groups (e.g., neighboring school districts only vs. entire NECTA division), weighting by schools’ population size, smoothing/rolling average case rates, and adjustments for various covariates, including measures of community burden of COVID-19 (**Table S2**). Our final analysis corrected for non-reporting as zeroes, considered all school districts within the NECTA division as comparators, and was weighted by school population size to capture the population impact of masking policies across our included school districts. Our main analysis did not adjust for measures of community level COVID-19 burden, as a growing body of evidence suggests schools as a driver of COVID-19 community burden,^38,39,62^ making it a mediating factor along the causal pathway rather than a confounder of the relationship between school masking policies and COVID-19 among students and staff.

### Descriptive Analysis

Finally, to assess whether school masking policies were enacted with attention to health equity, we compared timing of lifting/sustaining school masking policies across various school district characteristics, including student and sociodemographics and physical characteristics of the learning environment using data sources described above.

## RESULTS

Out of the 72 included school districts in the Boston-Newton-Cambridge NECTA division, only Boston and Chelsea Public Schools maintained masking throughout the study period (**Figure 1A**). Most school districts (n=46, 64%) lifted masking requirements when the statewide mask requirement was rescinded on February 28, 2022 (**Figure 1B**). The remaining 24 districts removed masking requirements in the following reporting week (n=17, 24%) or two weeks (n=7, 10%) after the statewide mandate was lifted.

**Figure 1.**
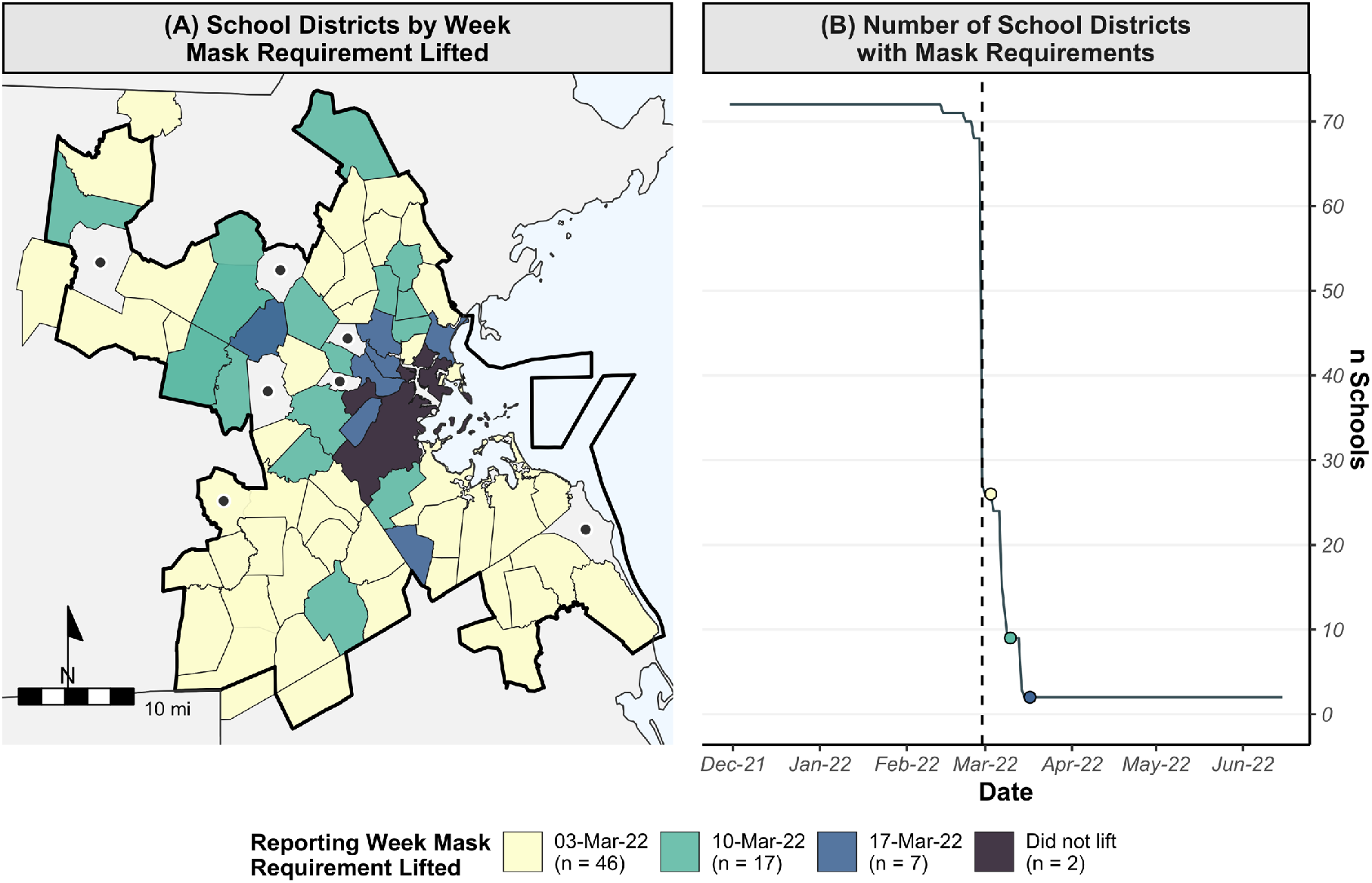
(A) Map of school districts in Boston-Cambridge-Newton NECTA division by reporting week in which mask requirements were removed and (B) number of school districts with mask requirements remaining in place over time^1^

Prior to the lifting of masking requirements, as cases declined during the initial BA.1 Omicron wave, reported COVID-19 case rates among students and staff were similar in Boston/Chelsea to school districts within the study area that later lifted their masking requirements (**Figure 2A**). However, following the lifting of masking requirements in late February through early March, reported COVID-19 rates diverged and were substantially higher in school districts that lifted their masking requirements compared to those observed in Boston and Chelsea Public Schools where mask requirements were sustained (**Figure 2A**). These trends held both overall (**Figure 2A**) and among students (**Figure 2B**) and staff (**Figure 2C**) separately.

**Figure 2.**
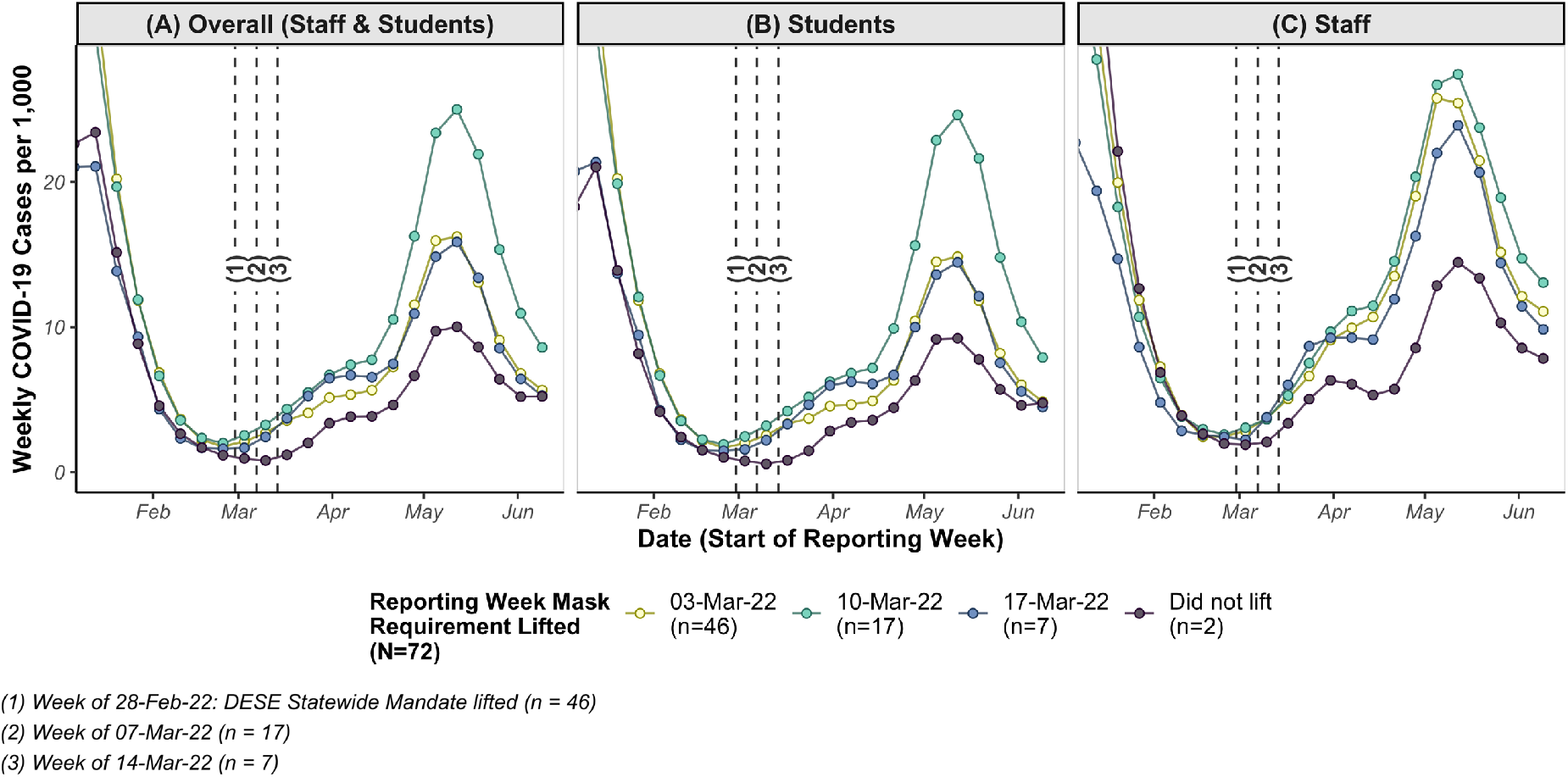
Weekly reported rate of COVID-19 cases (A) overall, (B) among students, and (C) among staff in Boston/Chelsea Public Schools, by week masking requirements were lifted for school districts within the Boston-Cambridge-Newton NECTA division.^2^

Next, we report individual weekly effects (**Figure 3A**) and cumulative effects over the study period (**Figure 3B**) from our difference-in-differences analysis. We did not observe meaningful patterns in differences between school districts in the period prior to the removal of the statewide school masking requirement. Despite substantial variance between schools in the weeks corresponding to the initial Omicron wave, the districts followed approximately the same trajectories during the surge with no clear patterns that might violate the parallel trends assumption (**Figure 3A.1**). In contrast, once masking requirements were lifted, we observed consistently higher case rates in those school districts compared to districts which continued masking requirements. Lifting of masking requirements was associated with significant increases in reported COVID-19 cases among students and staff in 12 out of 15 individual weeks in the post-period (**Figure 3A.1**). These impacts increased with the amount of time since a school district had lifted their mask requirement. In the first week following a lifting of masking requirements, we estimated an additional +1.44 (95% CI: +0.58, +2.29) COVID-19 cases per 1,000 students and staff, relative to school districts that had not yet lifted masking requirements. By the 9th week after a school district had lifted mask requirements, +9.68 per 1,000 (+7.11, +12.25) excess COVID-19 cases compared to school districts that had not yet lifted requirements (**Figure 3A.1**). We observed similar trends between students and staff, with slightly stronger weekly effects observed among staff (**Figure 3A.2**) compared to students (**Figure 3A.3**).

**Figure 3.**
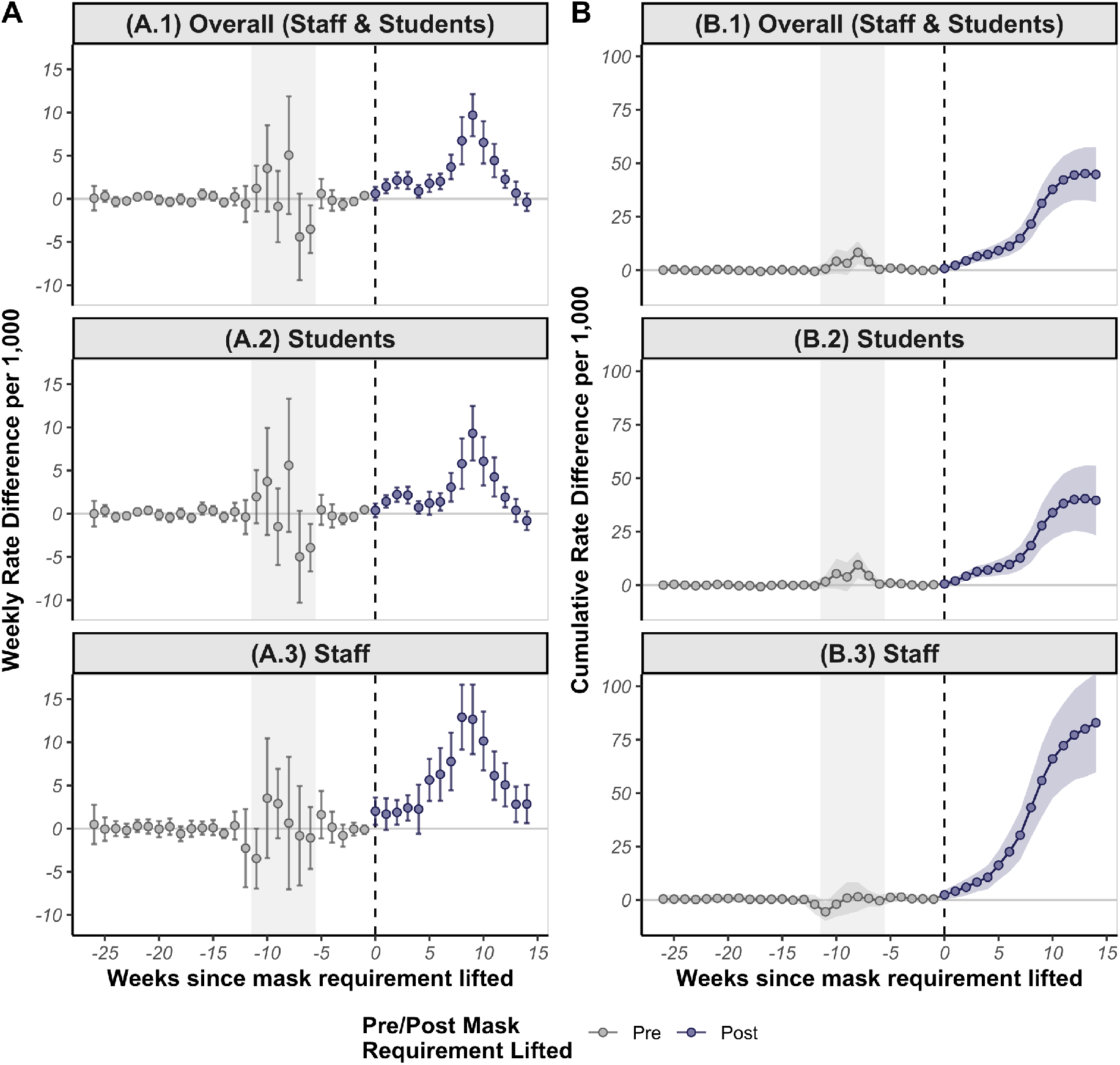
Weekly (A) and cumulative (B) differences in rate of COVID-19 overall (top), among students (middle) and among staff (bottom) in school districts that lifted masking requirements compared to school districts that had not yet lifted their masking requirements in that week.^3^

Importantly, the strength of the association between school masking requirements and COVID-19 case rates varied with the background rate of COVID-19 in the surrounding community, such that the strongest effects were observed in the weeks when background community case rates were at their highest for a given school district (**Figure S3, Figure S4**). Determining whether this is due to high community rates contributing to greater risk in unmasked schools or whether transmission in unmasked schools is contributing to higher community rates is difficult and beyond the scope of this paper. Nonetheless, these data clearly support a link between in-school transmission and community COVID-19 rates.

In the 15 weeks following the lifting of the statewide masking requirement in schools, the cumulative impact in school districts that lifted requirements increased over time for staff and students combined (**Figure 3B.1**), and for students (**Figure 3B.2**) and staff (**Figure 3B.3**) separately. Overall, we estimate that lifting of masking requirements was associated with an additional 44.9 (95% CI: 32.6, 57.1) cases per 1,000 students and staff over the 15 weeks since the lifting of the statewide mandate (**Table 1**). This excess rate corresponded to 11,901 (95% CI: 8,651, 15,151) total cases, representing 33.4% (95% CI: 24.3%, 42.5%) of cases in school districts that lifted masking requirements and 29.4% (95% CI: 21.4%, 37.5%) of cases in all school districts during that period.

**Table 1.**
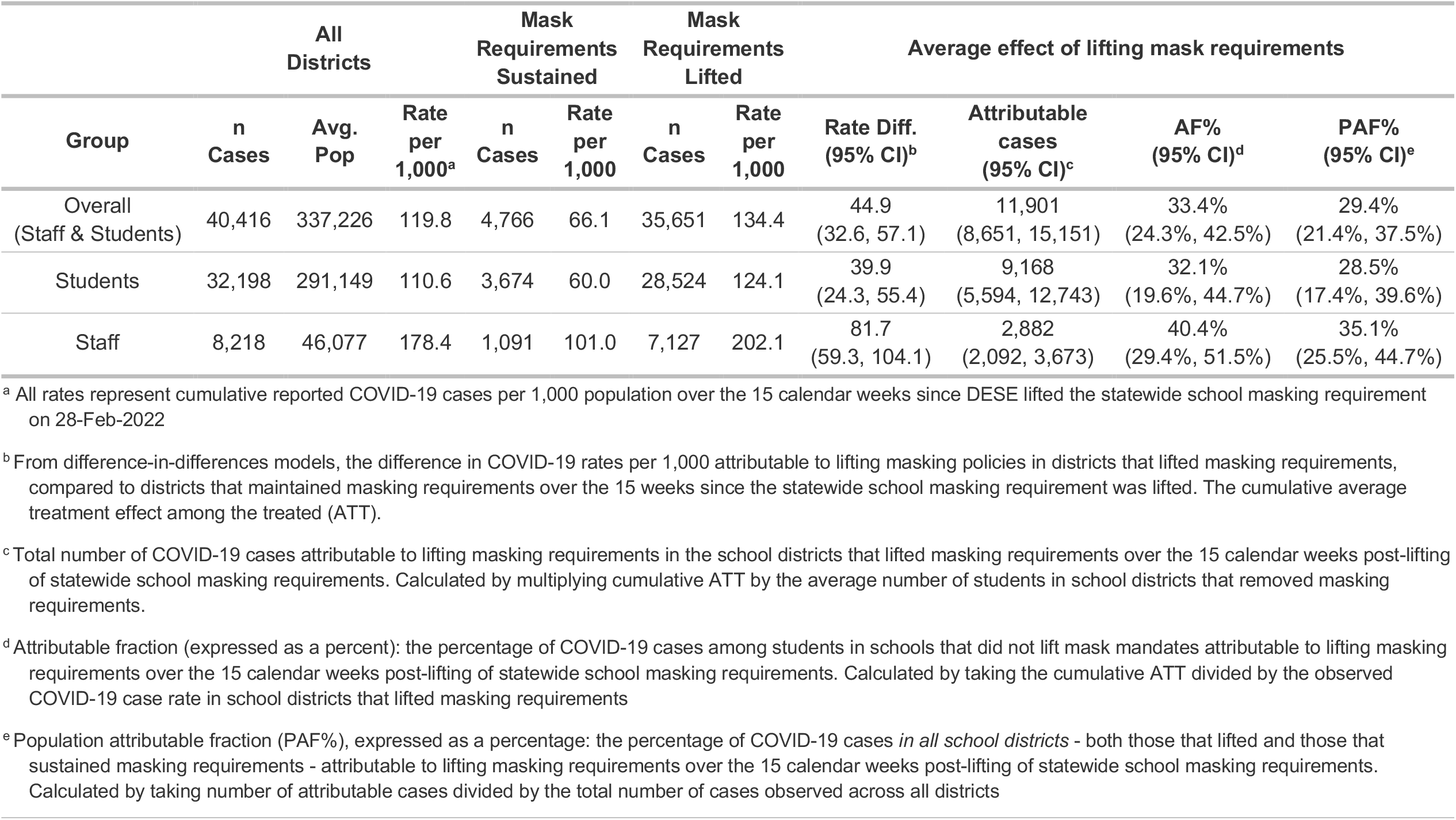
Rates of COVID-19 overall and by school district masking policies and estimated impact of lifting masking requirements in the 15 calendar weeks following lifting of statewide school masking requirements, overall and for staff and students separately

| Group | All Districts |  |  | Mask Requirements Sustained |  | Mask Requirements Lifted |  | Average effect of lifting mask requirements |  |  |  |
| --- | --- | --- | --- | --- | --- | --- | --- | --- | --- | --- | --- |
|  | n Cases | Avg. Pop | Rate per 1,000<sup>a</sup> | n Cases | Rate per 1,000 | n Cases | Rate per 1,000 | Rate Diff. (95% CI)<sup>b</sup> | Attributable cases (95% CI)<sup>c</sup> | AF% (95% CI)<sup>d</sup> | PAF% (95% CI)<sup>e</sup> |
| Overall (Staff & Students) | 40,416 | 337,226 | 119.8 | 4,766 | 66.1 | 35,651 | 134.4 | 44.9<br>(32.6, 57.1) | 11,901<br>(8,651, 15,151) | 33.4%<br>(24.3%, 42.5%) | 29.4%<br>(21.4%, 37.5%) |
| Students | 32,198 | 291,149 | 110.6 | 3,674 | 60.0 | 28,524 | 124.1 | 39.9<br>(24.3, 55.4) | 9,168<br>(5,594, 12,743) | 32.1%<br>(19.6%, 44.7%) | 28.5%<br>(17.4%, 39.6%) |
| Staff | 8,218 | 46,077 | 178.4 | 1,091 | 101.0 | 7,127 | 202.1 | 81.7<br>(59.3, 104.1) | 2,882<br>(2,092, 3,673) | 40.4%<br>(29.4%, 51.5%) | 35.1%<br>(25.5%, 44.7%) |

<sup>a</sup> All rates represent cumulative reported COVID-19 cases per 1,000 population over the 15 calendar weeks since DESE lifted the statewide school masking requirement on 28-Feb-2022
<sup>b</sup> From difference-in-differences models, the difference in COVID-19 rates per 1,000 attributable to lifting masking policies in districts that lifted masking requirements, compared to districts that maintained masking requirements over the 15 weeks since the statewide school masking requirement was lifted. The cumulative average treatment effect among the treated (ATT).
<sup>c</sup> Total number of COVID-19 cases attributable to lifting masking requirements in the school districts that lifted masking requirements over the 15 calendar weeks post-lifting of statewide school masking requirements. Calculated by multiplying cumulative ATT by the average number of students in school districts that removed masking requirements.
<sup>d</sup> Attributable fraction (expressed as a percent): the percentage of COVID-19 cases among students in schools that did not lift mask mandates attributable to lifting masking requirements over the 15 calendar weeks post-lifting of statewide school masking requirements. Calculated by taking the cumulative ATT divided by the observed COVID-19 case rate in school districts that lifted masking requirements
<sup>e</sup> Population attributable fraction (PAF%), expressed as a percentage: the percentage of COVID-19 cases *in all school districts* - both those that lifted and those that sustained masking requirements - attributable to lifting masking requirements over the 15 calendar weeks post-lifting of statewide school masking requirements. Calculated by taking number of attributable cases divided by the total number of cases observed across all districts

This effect was even more pronounced among staff, where lifting requirements was associated with an additional 81.7 (95% CI: 59.3, 104.1) COVID-19 cases per 1,000 staff over 15 weeks, representing 40.4% (95% CI: 29.4%, 51.5%) of all cases observed among staff in school districts that lifted masking requirements and 35.1% (95% CI: 25.5%, 44.7%) of all cases observed among staff across all school districts over this period. In sensitivity analyses, the results were robust to modeling specifications, including data cleaning measures, population weighting, restricting comparison group school districts to only neighboring school districts, and to adjusting for covariates including measures of community burden of COVID-19 (**Figure S5**).

Finally, we found that school district characteristics varied with the length of time they sustained their individual masking requirements following the lifting of the statewide mandate. We found that school districts that sustained masking protections for longer periods were those with a higher percentage of low-income students, students with disabilities, and English-learning students (**Figure 4A**) and a higher percentage of Black and Latinx students (**Figure 4B**) and staff (**Figure 4C**). In addition, school districts that sustained masking requirements for longer periods also had school buildings that were older, in poorer physical condition (a category that included ventilation/HVAC), and higher numbers of students per classroom (**Figure 4D**). In contrast, we found that school districts that lifted masking requirements earlier tended to have a lower percentage of low income and English-learning students, students with disabilities, Black and Latinx students and staff, fewer students per classroom, and newer buildings that were generally in better condition. These differences between districts have important implications for our results, as it clarifies that the increased COVID-19 rates in schools which removed masks are unlikely to be related to a higher baseline risk of SARS-CoV-2 exposure for students and staff outside of school. It also suggests that, despite newer buildings in better condition, ventilation in schools which lifted mask requirements was generally not sufficient to substantially impact SARS-CoV-2 transmission

**Figure 4.**
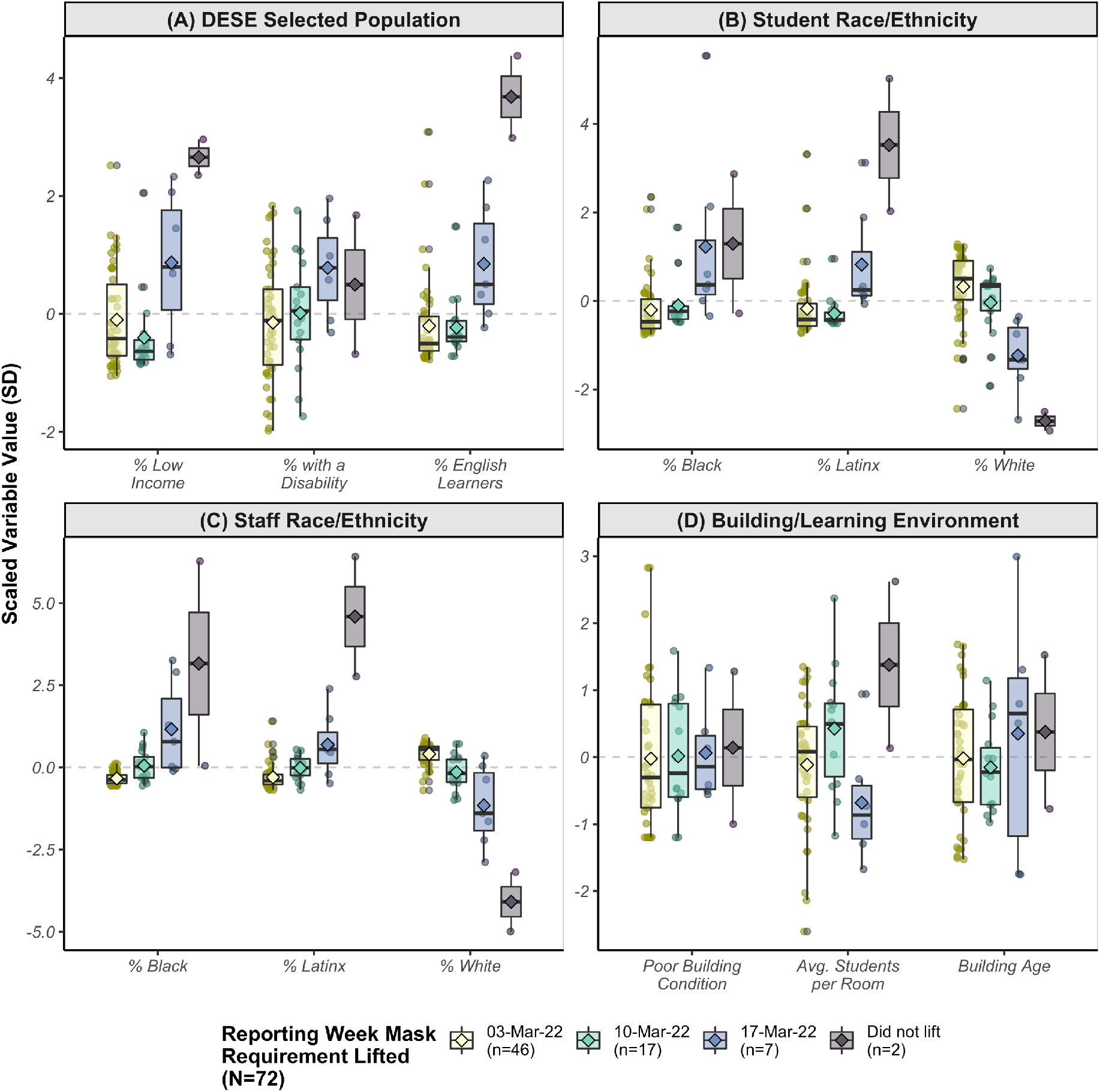
Distribution^4^ of selected characteristics of school districts by week masking requirements were lifted, including (A) percentage of students in selected populations as defined by the Massachusetts Department of Elementary and Secondary Education (DESE), race/ethnicity of (B) students and (C) staff, and (D) physical building conditions and learning environment characteristics from the 2016-2017 Massachusetts School Building Authority (MSBA) school survey

## DISCUSSION

Given the dynamic nature of the pandemic, it is critical to examine in near real-time the need for and impact of preventive measures, including masking in school settings. Schools are, and will continue to be, an important yet politically contested space in the COVID-19 response, making analyses such as this one particularly relevant to decision-makers. Our analysis adds to a growing body of literature documenting the benefits of universal masking policies in public schools during a period of highly transmissible SARS-CoV-2 variants and a rapidly changing COVID-19 policy environment.

Our paper documents significantly higher rates of reported COVID-19 in school districts that lifted masking requirements compared to those that sustained masking requirements following the rescinding of the Massachusetts statewide school masking order. Specifically, we estimate that rescinding mask requirements in school districts in Eastern Massachusetts during March 2022 may have contributed an additional 45 per 1,000 COVID-19 cases among students and staff in the 15 weeks following the end of school-based masking requirements. In total, this represents more than 9,000 cases among students and nearly 3,000 cases among staff. The Massachusetts Department of Elementary and Secondary Education requires that children who test positive for COVID-19 isolate for a minimum of 5 days, or until symptoms abate. In the best-case scenario, our results translate to a minimum of 17,505 days of school absence due to mandatory COVID-19 isolation in school children, and 6,547 days of teacher absence over the 15 weeks since the lifting of the statewide mandate (see methods in **Supplementary Appendix, Table S3**). Importantly, we observed the greatest impact of masking requirements in weeks with highest background community rates of COVID-19, underscoring the importance of early implementation and sustaining of school masking policies prior to and throughout surges. In addition, given the evolving understanding of the impact of long-COVID on children,^10–13^ our results suggest that masking requirements may be an important tool for school administrators and elected officials to consider as they plan for the upcoming school year.

Understanding COVID-19 policy decisions requires attention to power and existing historical and sociopolitical context.^23,63–65^ In the present study, we noted systematic differences in school masking policy choices by school district characteristics. Specifically, school districts that sustained masking requirements for longer periods tended to have school buildings in worse physical condition, more crowded classrooms, and a higher proportion of staff and students made vulnerable by historic and contemporary systems of oppression (e.g., racism, capitalism, xenophobia, and ableism). In addition, the only two school districts to sustain school masking requirements through June – Boston and neighboring Chelsea Public Schools – were also among the cities and towns in Massachusetts that have been most impacted by the COVID-19 pandemic to date. These differences in length of school masking policies may reflect an understanding among elected officials of how public policies are a key mechanism by which structural racism operates to produce health inequities.^23,30,33,63,65^ Structural racism and racial capitalism are fundamental causes of COVID-19 inequities.^22,23^ These forces operate via diverse mechanisms such as household crowding, employment in essential industries, and access to testing, vaccines and treatment, and differentially concentrate risk for both SARS-CoV-2 exposure and severe COVID-19 outcomes among low income and Black, Latinx, and Indigenous communities.^22–24,37,66^ Knowledge of these differential conditions may influence support/opposition to COVID-19 mitigation measures and policies in schools, including school masking.^67,68^ A growing body of work suggests that knowledge of these inequities may result in increasing support for protective measures among those directly impacted by structural racism and other systems of oppression while simultaneously decreasing support among systematically advantaged groups whose relative position largely insulates them from COVID-19 harms.^67–69^ In several studies and polls, Black and Latinx parents were less likely to have confidence that schools could reopen safely without additional protections and more likely to support school masking requirements.^67,68,70,71^ In contrast, when randomized to receive information about pervasive racial/ethnic COVID-19 inequities, white individuals were less likely to report concern about COVID-19, empathy for those vulnerable to COVID-19, and less likely to support COVID-19 prevention policies.^69^ Given vastly unequal environmental conditions shaping COVID-19b risk, universal school masking policies are an important measure for mitigating the impacts of structural racism and may be especially important in settings where other preventive measures such as upgraded ventilation/filtration may be more resource- and time-intensive to implement. School districts and policymakers should consider these inequitable impacts when making plans about masking policies for the upcoming school year.

Because universal school masking policies to prevent SARS-CoV-2 transmission has been a contentious issue, we anticipate a number of critiques which we address here, several of which are commonly levied at any study of masking in schools. One common critique of this type is that COVID-19 is rare and mild in children. However, we observed weekly COVID-19 case rates exceeding 20 per 1,000 students at times during the study. These high rates likely represent substantial educational disruptions and present increased risk of long-term complications (i.e., long-COVID). In addition, we observed larger benefits of sustained masking among staff, who may be at increased risk of severe COVID-19 outcomes. A second common critique of mandatory masking in schools is that alternative approaches to reducing transmission, such as improved ventilation, exist. While this may be true in theory, our findings make clear that the ventilation systems in Eastern Massachusetts school districts were insufficient to prevent all COVID-19 cases in schools. Indeed, school districts with newer buildings and better ventilation were more likely to remove masking requirements earlier, and thus our findings of increased COVID-19 incidence in the absence of masks demonstrate that ventilation likely remained insufficient in most schools in our sample, reinforcing the need for layered mitigation measures.

A key strength of this study is our use of staggered dates of removal of mask requirements, and difference-in-differences methodology. These approaches enabled us to estimate the impact of masking requirements despite differences between school districts. While it is true that there are several factors which differ between school districts, and which are related to SARS-CoV-2 exposure risk, difference-in-differences methodology is immune to those sources of confounding whenever they do not experience changes over time coinciding with the policy change of interest (e.g., differences in sociodemographics or vaccination rates). Furthermore, when we investigated the distribution of known COVID-19 risk factors between school districts, we found that those districts which removed masks were those which *a priori* would have been expected to have lower COVID-19 rates. This suggests that any residual confounding of our results by COVID-19 risk would have led to our analysis underestimating the harms of removing universal masking policies.

A key question in any study looking at schools is how background community COVID-19 rates interact with school-specific rates. There are two potential reasons why community levels may be relevant, and the correct analytic decision on whether to include community levels as confounder or not, is not straightforward. In sensitivity analyses, we found that the benefits of universal masking requirements persisted even after controlling for several measures of community COVID-19 incidence. We, however, did not prioritize these results in our main analyses, as we argue it is more appropriate to consider community rates of COVID-19 as part of the causal effect of school masking policies rather than a source of bias. School and community COVID-19 levels are so closely linked that it is difficult to rule out at least some of the variation in community COVID-19 rates being a direct consequence of changing school case rates.^63^ That is, students and staff can infect their family and community members and vice versa. In addition, while some community-level policies changed during Spring 2022, these changes did not perfectly coincide with the school mask mandates being rescinded, nor did the community mask policies always align with school mask policies. Overall, we anticipate that although community case rates change over time and are connected to school case rates, the ways in which community cases and school cases are related should not depend on school masking policies.

A second reason to consider community COVID-19 cases is that there may be spillover between school districts or communities as individuals move between areas. This is not an issue of confounding, but rather a potential threat to the validity of estimating causal effects of COVID-19 prevention policies. Spillover between communities and school districts may reduce the difference in COVID-19 rates and has the potential to alter the case growth trajectory in masked schools. This latter issue is a potential threat to the parallel trends assumption required for difference-in-differences analyses. To address this, we used the staggered policy adoption model of difference-in-differences.^59–62^ This approach uses all pre-policy change data as unexposed data, and aligns the comparisons for school districts which unmask to contemporaneous control districts, allowing us to estimate the impact of removing masks, uncoupled from calendar time. As a result, we anticipate that any remaining impact of spillover on our findings would be to decrease the estimated impact of removing masks, making our results an underestimate of the harms caused by removing mask mandates earlier.

Overall, our findings should be interpreted as the impact of universal masking policies, not masks *per se*, given that it is unlikely that all children and families removed masks when the requirement was removed, as masks were still encouraged in most school settings and utilized by many. Despite this, the impact of lifting required masking policies was substantial.

## CONCLUSION

Undoubtedly, the Omicron wave will not be the final COVID-19 surge, and ongoing efforts to minimize the impacts of COVID-19 in school settings, including evidence-based public policy, are urgently needed. Our results underscore the importance of centering health equity in these policy choices and support universal school masking policies as an important piece of a layered mitigation strategy to reduce COVID-19 risk among those made vulnerable by structural racism and other systems of oppression. If trends from prior years persist, surges may be especially likely to occur in late December and January. Many students and staff will likely have spent time traveling, gathering indoors, or in other high-risk transmission settings during winter break, and epidemiologic best practices suggest a 14-day quarantine following these types of activities to prevent onward transmission. We recommend that school districts develop mitigation plans proactively in anticipation of a winter COVID-19 wave during the 2022-23 school year. In particular, requiring masks in schools in December and January, with a clear *a priori* decision threshold for removing masks in March or later as the winter wave abates, could be an effective strategy to minimize the impact of COVID-19 in school settings.

## Data Availability

All data utilized in this study are publicly available through the Massachusetts Department of Elementary and Secondary Education (https://www.doe.mass.edu/covid19/positive-cases/default.html#weekly-report); Massachusetts Department of Public Health (https://www.mass.gov/info-details/covid-19-response-reporting); and the Massachusetts School Building Authority (https://www.massschoolbuildings.org/programs/school_survey)

## Footnotes

1 Black dots denote the n=7 school districts excluded from the analysis due to unreliable data reporting, see **Figure S1**

2 Dates on the x-axis were restricted to the period immediately before and after universal school masking requirements were lifted statewide and in most school districts. Difference-in-differences analysis includes all weeks in the 2021-2022 school year.

3 *Results are modeled estimates from our difference-in-differences analysis. The grey band in the background of the plot depicts the initial BA*.*1 Omicron wave in December 2021-January 2022*.

4 Variables are scaled to enable them to be depicted on the same scale – zero represents the mean value with units in standard deviations

## Notes

**Conflict of Interest Statement.** All authors have completed the ICMJE uniform disclosure form at www.icmje.org/coi_disclosure.pdf and declare no competing interests.

### Competing Interest Statement

The authors have declared no competing interest.

### Funding Statement

This study did not receive any external funding.

