## Supplementary Appendix for "Impact of Lifting School Masking Requirements on Incidence of COVID-19 among Staff and Students in Greater-Boston Area School Districts: A Difference-in-Differences Analysis"

### SUPPLEMENTARY METHODS

*Inclusion/exclusion criteria*

As summarized in **Table S1** below, in total, N=400 districts reported COVID-19 data to DESE. After excluding n=211 charter, vocational, and technical school districts and restricting to school districts within the Boston-Newton-Cambridge New England City and Town Area (NECTA), n=79 schools remained (See **Figure S1**). We additionally excluded n=7 school districts with unreliable data for a final sample size of n=72 school districts (See **Table S1)**.

A school districts was considered unreliable if it had either:

1. >10 weeks with zero reported cases among students and staff

   ***<u>OR</u>***

2. More than 5 reporting weeks with either:
   a. Zero reported cases among students and staff and at least 1 positive pool test

      ***<u>OR</u>***

   b. Zero cases reported among students and staff and >10 cases reported on average over the past 4 weeks

*Sensitivity analyses*

We conducted several sensitivity analyses to assess whether our results were robust to various data cleaning steps and model specifications. Our sensitivity analyses fell into five broad categories: (A) data cleaning; (B) population weighting; (C) smoothing; (D) modified control groups; and (E) adjustment for covariates. For each of these categories, additional details of these analyses are described in **Table S2**, below. Results of the sensitivity analyses described below can be found in **Figure S5**.

*Calculation of school days missed (presented in discussion)*

To calculate the number of missed school days attributable to lifting of mask requirements presented in the discussion, we started with the estimated number of cases attributable to lifting of the masking requirements (presented in **Table 1** of main results). We multiplied the attributable cases by the minimum 5-day isolation requirement set by DESE during our study period. We then multiplied this product by the proportion of calendar days that children were in school to account for the fact that not all isolation days after a COVID-19 case is identified would fall on school days (i.e., school holidays, weekends, etc.). To get the proportion of calendar days during our study we used the 2021-2022 Boston Public Schools calendar[1] and determined that there were 181.5 school days out of 290 calendar days and therefore in total, 62.6% of calendar days of the school year are days students/staff spend in schools. The results of these calculations are shown in **Table S3** below.

Finally, to reflect the statistical uncertainty in our estimates, we present the lower confidence interval for the estimated missed number of school days, as this represents a conservative estimate based on our models (e.g., "...our results translate to a minimum of 17,505 days of school absence due to mandatory COVID-19 isolation in school children"). The numbers we present in our discussion are highlighted and underlined in **Table S3**.

[1] 2021-2022 Boston Public Schools Calendar, available: https://www.bostonpublicschools.org/cms/lib/MA01906464/Centricity/Domain/4/BPS%20Cal%20SY22_FINAL.pdf

**SUPPLEMENTARY TABLES & FIGURES**

**Figure S1.** *Map of Massachusetts school districts for inclusion in the present study. Black dots indicate the n=7 school districts[2] within the Boston-Cambridge-Newton NECTA division that were excluded from the study due to unreliable data.*

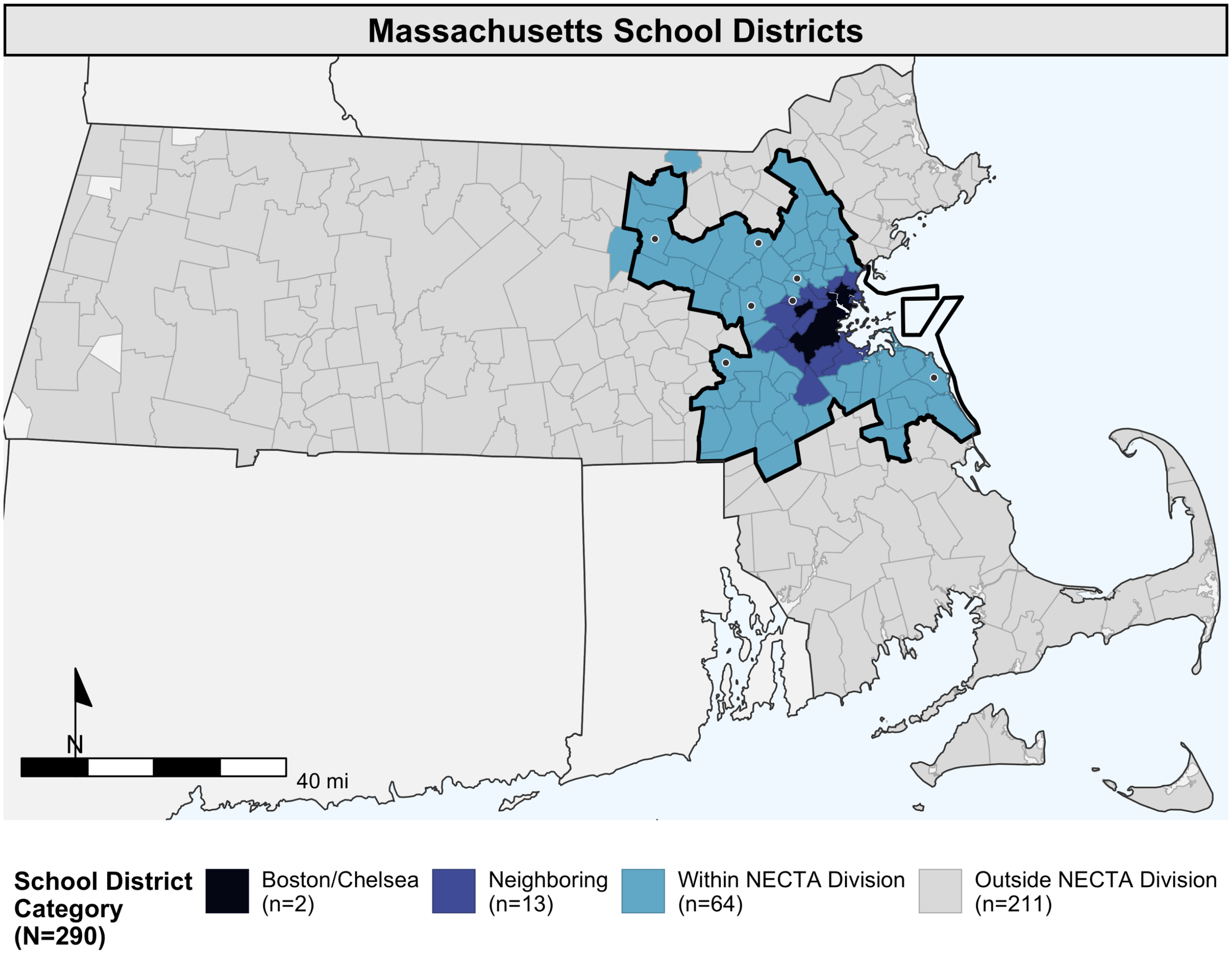

[2] Excluded school districts: Arlington, Bedford, Harvard, Scituate, Sherborn, Weston, and Watertown

**Table S1.** *Inclusion and exclusion steps and criteria and included/excluded school districts in final sample*

| n Remaining | n Excluded | Exclusion Step |
|---|---|---|
| 400 | | Total school districts reporting COVID-19 data to DESE |
| 79 | (-211) | Exclude charter, vocational, and technical school districts |
| 72 | (-7)* | **Final sample;** Exclude unreliable districts; districts were excluded if either:<br>1. School district had >10 weeks with 0 cases reported among students and staff<br>**OR**<br>2. School district has >5 weeks where either:<br>a. 0 cases reported among students and staff, but district reported at least 1 positive pool test<br>**OR**<br>b. 0 cases reported among students and staff and ≥10 cases reported on average over the past 4 weeks |
| ***Excluded Districts in NECTA (n=7):** Arlington, Bedford, Harvard, Scituate, Sherborn, Weston, Watertown<br><br>**Included districts:** Andover, Belmont, Boston, Braintree, Brookline, Burlington, Cambridge, Canton, Carlisle, Chelsea, Cohasset, Concord, Dedham, Dover, Everett, Foxborough, Franklin, Hanover, Hingham, Holbrook, Hull, Lexington, Lincoln, Lynnfield, Malden, Mansfield, Marshfield, Maynard, Medfield, Medford, Medway, Melrose, Millis, Milton, Needham, Newton, Norfolk, North Reading, Norwell, Norwood, Quincy, Randolph, Reading, Revere, Rockland, Saugus, Sharon, Somerville, Stoneham, Stoughton, Sudbury, Wakefield, Walpole, Waltham, Wayland, Wellesley, Westwood, Weymouth, Wilmington, Winchester, Winthrop, Woburn, Wrentham, Acton-Boxborough, Ayer Shirley School District, Concord-Carlisle, Dover-Sherborn, Groton-Dunstable, King Philip, Lincoln-Sudbury, Nashoba, Whitman-Hanson | | |

**Table S2.** *Descriptions of various sensitivity analyses for difference-in-differences analyses. The descriptions here correspond to the results presented in* ***Figure S5****.*

<table>
<tr><th>Sensitivity Analysis Category</th><th>Type & Details</th></tr>
<tr><td rowspan="4">(A) Data Cleaning Steps<br><br>Data cleaning steps address two main challenges – 2-week reporting weeks and missing data reported as zeros.<br>● For n=4 reporting weeks (corresponding to the weeks after Thanksgiving recess, winter recess, February recess, and spring recess), DESE asked that school districts report all cases for the prior two weeks (i.e., reporting period was 2 calendar weeks rather than 1 calendar week).<br>● DESE data does not distinguish between zero cases and non-reporting. Upon examining the data, we noted that often school districts would account for a zero report in the prior week by reporting two weeks' worth of cases in the following week (i.e., school districts were over-correcting for not reporting in the following week).</td>
<td>Raw Data: data as reported by DESE with no changes<br>● 2-week reporting weeks: considered data missing for first week, rate calculated over 14 person-days for 2nd week<br>● Zero reporting weeks: Considered true zeros/not corrected</td></tr>
<tr><td>Correct for 2-week reporting periods: raw data from DESE, but corrects for 2-week reporting weeks<br>● 2-week reporting weeks: assumes equal rate across both weeks<br>● Zero reporting weeks: Considered true zeros/not corrected</td></tr>
<tr><td>Remove non-reporting zeros: Corrects for 2-week reporting periods (as above), and excludes some zeros as non-reporting weeks<br>● 2-week reporting weeks: assumes equal rate across both weeks<br>● Zero reporting weeks: Exclude reporting weeks with 0 reported cases across students and staff if:<br>○ 0 Cases reported, but at least 1 positive pool testing OR<br>○ 0 cases reported and >10 cases reported on average over the past 4 weeks</td></tr>
<tr><td>Correct non-reporting zeros: Corrects for 2-week reporting periods (as above), considers some zeros as non-reporting, and assumes over correction in the following week<br>● 2-week reporting weeks: assumes equal rate across both weeks<br>● Zero reporting weeks: Exclude reporting weeks with 0 reported cases across students and staff with same rules as above; additionally assume that schools correct for this by reporting prior cases in the following week (i.e., rate in week following zero reporting week is calculated for 2 weeks of person-days)</td></tr>
</table>

| Sensitivity Analysis Category | Type & Details |
|---|---|
| **(B) Population Weighted**<br><br>*Traditional difference-in-differences models treat all observations (i.e., school districts) as having equal weight. Weighting accounts for large variation in school districts size and measures the population-level impact of lifting of masking requirements.* | **Population weighted:** Weekly school district observations are weighted by the school district population size in difference-in-difference models |
| **(C) Smoothed**<br><br>*For similar reasons to (A) above, using rolling averages smooths over the data partially correcting for reporting zeroes instead of missing values.* | **3-week rolling average:** We used a 3-week centered rolling average to minimize noise in the data while preserving the timing of changes in trends (i.e., leading or lagging averages would have shifted the timing of changes in case rates) |
| **(D) Modified Control Group**<br><br>*Our main analysis considers all public school districts in the Boston-Newton-Cambridge NECTA. In these analyses, we considered different definitions for control counties as school districts more proximate to Boston/Chelsea may serve as better control groups than those further away* | **Primary and secondary neighbors:** Excludes school districts within NECTA that do not share borders Boston/Chelsea or Boston/Chelsea's neighboring school districts |
| | **Primary neighbors only:** Excludes school districts within NECTA that do not border Boston/Chelsea |
| **(E) Adjusted for city/town characteristics**<br><br>*Our main analysis did not adjust for any time-dependent covariates; however, we considered these covariates in sensitivity analyses in difference-in-differences models* | **City/Town Population:** *Adjusted for city or town population size in which the school district was located. Data from MA DPH (16-Jun-2022).* |
| | **Community COVID-19 % Test Positivity:** *Adjusted for city or town's total number of positive tests divided by the total number of tests over the past two weeks. Data from MA DPH (16-Jun-2022).* |
| | **Community COVID-19 Case Rate (Reported):** *Adjusted for city or town's total number of reported cases tests divided by the city/town population over the past two weeks. Data from MA DPH (16-Jun-2022).* |

| Sensitivity Analysis Category | Type & Details |
|---|---|
| | **<u>Community COVID-19 Case Rate (Corrected):</u>** City/towns' COVID-19 case rate adjusted for underreporting using the percent test positivity using methodology from:<br><br>Chiu WA, Ndeffo-Mbah ML. Using test positivity and reported case rates to estimate state-level COVID-19 prevalence and seroprevalence in the United States. *PLoS Comput Biol*. 2021;17(9): e1009374.<br><br>*Data from MA DPH (16-Jun-2022).* |
| | **<u>School district size:</u>** *Adjusted for school district population size (i.e., number of students/staff); data from DESE* |

**Figure S2.** *Weekly reported rate of COVID-19 cases (A) overall, (B) among students, and (C) among staff in Boston/Chelsea Public Schools, neighboring school districts and non-neighboring districts within the Boston-Cambridge-Newton NECTA division.*[3]

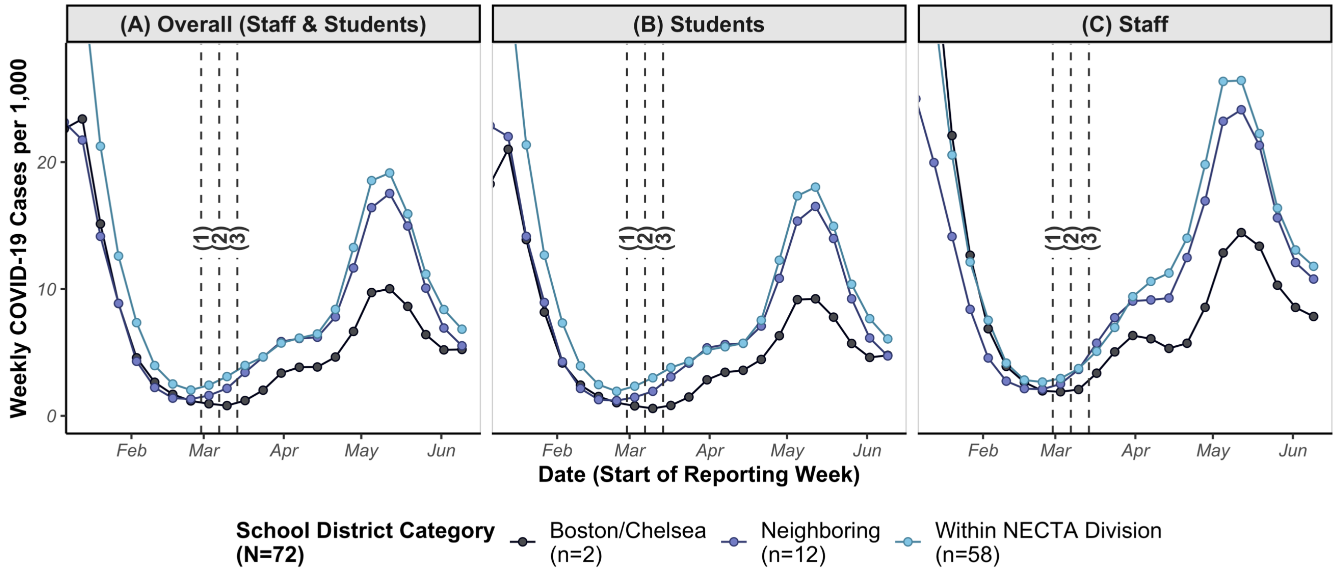

*(1) Week of 28-Feb-22: DESE Statewide Mandate lifted (n = 46)*
*(2) Week of 07-Mar-22 (n = 17)*
*(3) Week of 14-Mar-22 (n = 7)*

---

[3] Dates on the x-axis were restricted to the period immediately before and after mandates were lifted statewide and in most school districts. Difference-in-differences analysis includes all weeks in the 2021-2022 school year.

**Figure S3.** *Weekly differences in rate of COVID-19 among staff and students in school districts that lifted masking requirements compared to school districts that had not yet lifted their masking requirements colored by background community COVID-19 rate*[4]

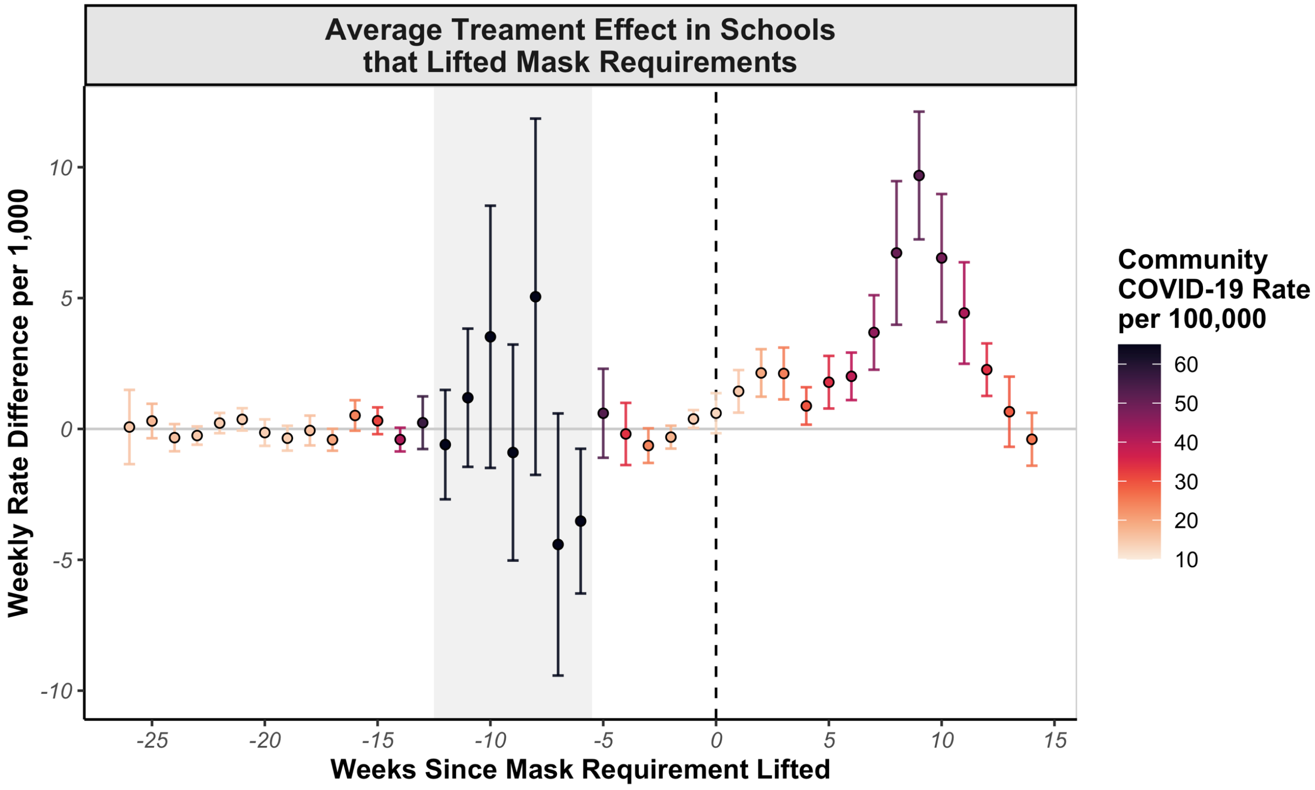

[4] Estimates are from difference in differences models and identical to those depicted in **Figure 3A.1** in the main text; Darker colors indicate higher background COVID-19 rates in the corresponding city/town; Grey band on the plot depicts the initial BA.1 Omicron wave in December 2021-January 2022

**Figure S4.** Relationship between COVID-19 case rates in surrounding communities (x-axis)[5] and weekly effect estimate sizes for the average treatment effect of removing masking requirements (y-axis) before (pre-period) and after (post-period) masking requirements were lifted

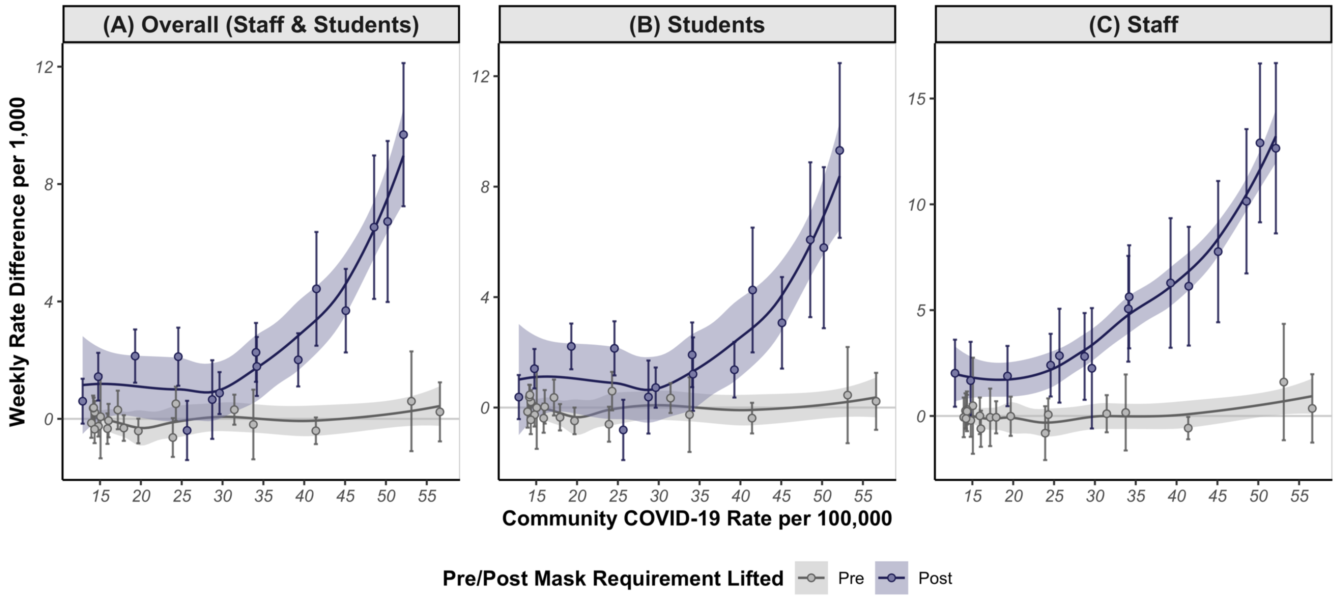

[5] X-axis attenuated at a weekly COVID-19 community case rate of 55 per 100,000 persons to show the range community case rates observed both in pre- and post- masking period (i.e., excluding highest Omicron peak values in early January 2022)

**Figure S5.** *Cumulative effect estimates for the impact of mask removal overall and for students and staff under various sensitivity analyses*[6] *including (A) varied data cleaning and suppression steps, (B) weighted by school district size (i.e., population),*[7] *(C) smoothed using 3-week rolling averages, (D) using varied control school districts, and (E) adjusting for various covariates (See methods section in Supplementary Appendix and* ***Table S2*** *for additional details)*

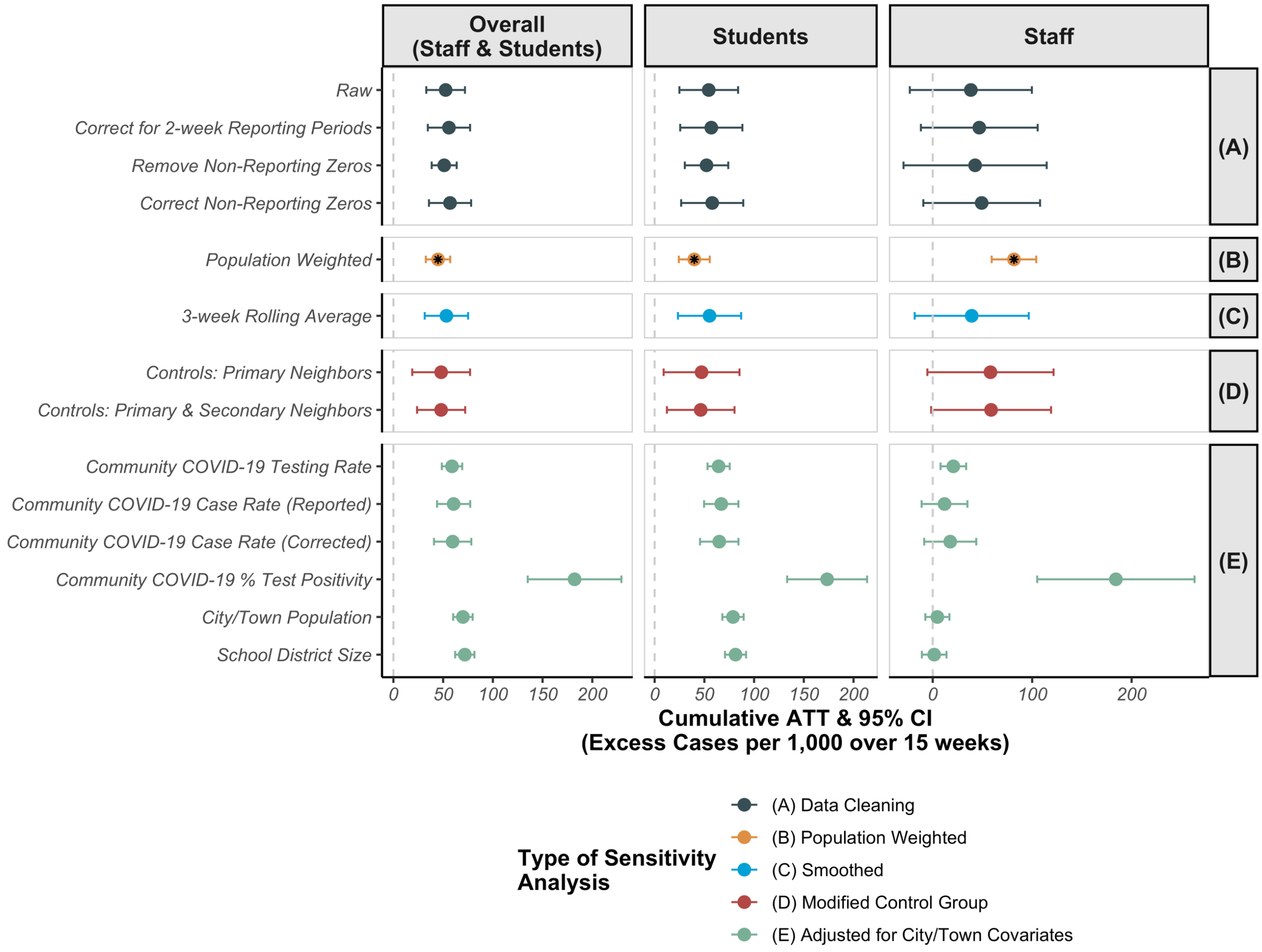

[6] These estimates correspond to estimates presented in **Table 1** of main text
[7] Black asterisk indicates the results presented in main analyses (i.e., results in main text are population weighted)

**Table S3.** *Calculation for number of missed school days following 5-day isolation for COVID-19 cases attributable to lifting school masking requirements (See* ***Supplementary Methods*** *for details of calculation)*

| | **Attributable COVID-19 Cases**[8] | | | **n Missed Days**[9] | | |
|---|---|---|---|---|---|---|
| | *Estimate* | *95% LCL* | *95% UCL* | *Estimate* | *95% LCL* | *95% UCL* |
| Overall (Staff & Students) | 11,901 | 8,651 | 15,151 | 37,242 | 27,072 | 47,412 |
| Staff | 2,882 | 2,092 | 3,673 | 9,019 | *6,547*[10] | 11,494 |
| Students | 9,168 | 5,594 | 12,743 | 28,690 | *17,505* | 39,877 |

---

[8] Estimates from difference-in-differences models presented in main text **Table 1**

[9] Calculated as: n Attributable Cases x 5 days isolation per case x 0.6259; where 0.6259 represents the proportion of calendar days in school (181.5 school days out of 290 calendar days)

[10] The highlighted and underlined numbers are presented in main text discussion section
